# To Include or Not to Include? A prescription from the pharmacy on how to use active learning assisted screening in systematic reviews

**DOI:** 10.1101/2025.09.26.25336705

**Authors:** Rinus G. Verdonschot, Tinne Dilles, Caitriona Cahir, Marjan De Graef, Renata Vesela Holis, Juliane Fryden, Petra Denig, Tamasine Grimes, Fatma Karapinar-Carkıt, Marieke Schor

## Abstract

**Background:** Systematic reviews are critical for evidence-based decision-making but require significant manual effort during the screening stage, which is labor-intensive and prone to error. Active learning (AL)-assisted screening tools have emerged to address these challenges. However, guidance for using AL-assisted screening in systematic reviews - especially those employing broad search strategies with heterogeneous results - is limited. This study aims to assess the effectiveness and reliability of AL-assisted screening for large, heterogeneous datasets. Specifically, it evaluates the comprehensiveness and necessity of the recommended SAFE procedure, examines the influence of different labeling strategies, and investigates whether AL-assisted screening can aid in reducing manual screening errors.

**Methods:** Screening of four large, heterogeneous datasets from medication management systematic reviews was simulated using ASReview. The datasets ranged from 3475 to 16218 records. For these datasets 0.08 to 1% of records were included in the final systematic review. Our simulations systematically varied all parameters defined by the SAFE procedure. Recall versus sampling behavior was analyzed, with a focus on the impact of parameter choices on retrieving records selected for full text inclusions and on reducing the number of records to be screened.

**Results:** AL-assisted screening can effectively reduce the number of records to screen by almost 90% without increasing the risk of missing relevant records in comparison to manual screening. For three of our datasets, the best performance (100% recall of full text includes and 89-90% reduction in the number of records to screen) is achieved when using the SAFE procedure in combination with the elas-u4 and elas-h3 models and full text labeling. This choice of parameters results in only 87% recall of full text includes for the remaining dataset (16218 records, 0.6% title/abstract includes, 0.08% full text includes). For this dataset, the best performance (100% recall, 90% screening reduction) is achieved when using the SAFE procedure with the simpler Naive Bayes model and TF-IDF feature extractor and title/abstract labeling.

**Conclusions:** AL-assisted screening can safely and effectively reduce the workload needed to screen the large, heterogeneous datasets common in medication management systematic reviews. We recommend the modified SAFE procedure using full-text labels and the elas models. If the estimated ratio of full text includes is very low, it may be more appropriate to use the original SAFE procedure with title/abstract labeling.

## 1 Introduction

Literature reviews aim to bring together all relevant available knowledge on a topic. Particularly reviews which are performed in a systematic and transparent way following standardized procedures, such as the systematic review, meta-analysis, and the scoping review, are now recognized across a wide variety of disciplines as essential tools for evidence-based decision making and identification of knowledge gaps [1–3].

However, the same rigorous methodology that makes these reviews valuable and trustworthy also makes them very labor-intensive. One of the most arduous steps in the review process is screening of the search results to identify the minority of records eligible for inclusion in the review. Screening is typically done in two stages. First the titles and abstracts of retrieved articles are compared to the criteria for inclusion and exclusion. Articles clearly not meeting the inclusion criteria and/or meeting one or more of the exclusion criteria are eliminated at this stage. Subsequently, the full text of articles which cannot confidently be excluded based on their title and abstract is assessed to come to a final set of articles to be included in the review.

Currently, full manual screening remains the gold standard in systematic reviews. However, this approach is still susceptible to errors, including the mistaken exclusion of articles during the title and abstract screening stage that should have been included in the review [4, 5]. Moreover, full manual screening is very labor intense.

Advances in large language models (LLMs) and machine learning have opened the way for the development of artificial intelligence (AI) based tools aimed at reducing the amount of manual work involved in the review process [6–8]. In particular tools leveraging active learning hold great promise to reduce the workload in screening [9]. Using active learning (AL)-assisted screening, the probability of inclusion for each article is automatically calculated based on a small initial training dataset provided by the researcher. The system then ranks articles based on the inclusion probability and presents articles to the researcher based on this ranking. With each decision made, the ranking is updated dynamically, allowing researchers to focus primarily on articles which are likely to be (most) relevant articles. This approach of AL-assisted screening has the potential to reduce the screening burden in terms of the number of records assessed by a human by as much as 90% [10–13].

The potential of AL-assisted tools to lower the workload associated with screening whilst maintaining a low error rate makes the use of these tools very attractive, particularly for larger datasets. For many reviews it is impossible to formulate a highly specific search strategy, for instance because the research question cannot be captured by well-defined keywords and/or controlled vocabulary terms. This is a common issue in fields like public health, health economics, and medication management, among others. To ensure sufficient sensitivity, search strategies for such reviews are kept broad and unspecific, often resulting in retrieval of very large numbers of search results with a high (*≥* 99%) proportion of irrelevant results [14].

Estimates of potential efficiency gains and recommendations of best practices developed for reviews with a specific search strategy may not generalize to those with broad, unspecific search strategies. The AL-assisted screening tools may struggle with the even larger imbalance between relevant and irrelevant sources in the dataset. Moreover, the search results retrieved with broad searches are likely to be more heterogeneous (e.g. in terminology used) than those obtained with more specific searches. This heterogeneity may also affect the performance of AL-assisted screening.

Despite the need for clear guidance on the use of AL-assisted screening in reviews with broad search strategies, there are currently only a limited number of studies evaluating the usefulness and reliability of AL-assisted screening for this type of dataset [15–19]. Although these all indicate that a reduction in screening workload can be obtained, they are diverse in their methodologies and outcomes, and do not provide clear guidance on best practices for researchers considering using AL-assisted screening.

When it comes to formulating guidance, there are several parameters which affect the reliability of AL-assisted screening, some *intrinsic* to the tool and some dependent on the user and thus *extrinsic* to the tool (see section 2.2). Intrinsic parameters are the feature extractor, the active learning model, the query strategy, and the balancing technique. Extrinsic parameters affecting the performance of AL-assisted screening are the selection of prior knowledge, the labeling strategy applied by the screeners, and the stopping criterion.

Several studies have evaluated the impact of one or more intrinsic or extrinsic parameters on the performance of AL-assisted screening [18, 20–26]. The most comprehensive advice on how to use AL-assisted screening such that the risk of missing out relevant records is minimized currently seems to be provided through the SAFE procedure [25]. This procedure provides advice on the optimal selection of all parameters with the exception of the labeling strategy applied by the screeners. It also provides a clear workflow for review authors to follow. The SAFE procedure has been applied in several recent reviews to speed up title-abstract screening [27, 28]. However, independent validation of the procedure based on existing datasets is lacking and guidance on the labeling strategy should be incorporated as a previous study has highlighted potential impact of noisy labeling [24].

Here, we aim to provide clear guidance on how best to use AL-assisted screening in reviews with broad search strategies based on a thorough validation study using four datasets from recent reviews in medication management for which highly specific search strategies could not be formulated without sacrificing sensitivity. The questions we aim to answer are:

- Is the SAFE procedure indeed sufficient and necessary to limit the risk of missing out on full text includes?
- Should crisp (full text) include labels be used or are noisy (title/abstract) labels sufficient to obtain the desired performance of AL-assisted screening?
- Can AL-assisted screening tools aid in reducing human error in screening?

## 2 Methods

### 2.1 Tools for AL-assisted screening

There are several tools available offering AL-assisted screening, for instance Rayyan, Abstrackr, ASReview, and Colandr [11, 29–31]. For recent comprehensive comparisons we refer to the works of Burgard et al. or Teijema et al. [9, 32]. We used ASReview [11, 33] for our validation. ASReview has the advantages that it is open source software, ensuring full transparency of the algorithms and methodologies behind any suggestions made by the software, and that it gives users full control of all intrinsic parameters.

### 2.2 Intrinsic and extrinsic parameters in AL-assisted screening

Regardless of the tool used for AL-assisted screening, choices for tool-intrinsic and tool-extrinsic parameters will need to be made.

The tool-intrinsic parameters are coded into the tool and can be selected from the settings. These include the feature extractor, the classifier, the query strategy, and the balancing technique. Any AL-assisted screening tool must first convert textual data into a numerical format it can process. This transformation is performed by a feature extractor. In this work we test the feature extractors term frequency - inverse document frequency (TF-IDF) with or without bigrams, sentence bidirectional encoder representations from transformers (sBERT), and mixed bread artificial intelligence embed large version 1 (mxBAI). Subsequently, this information will be used by a classifier to reorder the pile of records and put more relevant records to the top of the pile. Classifiers tested in this work are naive Bayes (NB), random forests (RF), and support vector machines (SVM). The ‘query strategy’ tells the model what the next thing is that should be shown to the user. Maximum, for example, means ‘show the record that is most likely to be relevant’. However, one could also choose 95% maximum and 5% random, i.e., to find records that might deviate from the other records but might still be relevant. All our simulation used maximum as a query strategy. Lastly, the ‘balancing technique’ pertains to how well a model can learn from imbalanced data. Review datasets are imbalanced as they typically contain many more irrelevant records than relevant records. Thus, a model would encounter many more irrelevant examples compared to relevant examples. The balancing approach aims to correct for this by weighing information from relevant and irrelevant records differently. All our simulations used balanced as balancing approach.

Tool-extrinsic parameters deal with how a human decides to interact with the AL-learning screening tool and include the selection of prior knowledge, the labeling strategy applied by the screeners, and the stopping criterion. The first step in AL-assisted screening consists of training the model by presenting it with a selection of prior knowledge (a.k.a. training data or “priors”). This prior knowledge allows the model to produce an initial ranking of unviewed records. Subsequent labeling decisions provide information to refine the model and update the ranking. How closely the labeling decisions reflect which sources will ultimately be included in the review therefore directly impacts the ranking and thereby the reliability with which relevant articles will surface. Finally, the risk of ending screening before all relevant articles have been identified should be limited by using appropriate stopping heuristics. The stopping heuristic outlines the criteria which should be met in order to terminate screening.

#### 2.2.1 SAFE Procedure

To date, the SAFE procedure seems the most comprehensive advice on how to use AL-assisted screening such that the risk of missing out relevant records is minimized [25]. It suggests choices for all tool-intrinsic parameters and all tool-extrinsic parameters except for the labeling strategy. The SAFE procedure specifies a four-stage process:

- Phase 1: Using a combination of a simple model and feature extractor (e.g. Naive Bayes with TF-IDF) setup screening using a random set of records (e.g. 1% of the dataset) as prior knowledge.
- Phase 2: Screen using active learning with the model and feature extractor of Phase 1 until a four-fold stopping heuristic is met.
- Phase 3: Use the output of phases 1 and 2 as prior knowledge to train a more advanced model and feature extractor (e.g. an ensemble learning method such as random forests with sBert as the feature extractor) and continue screening until a simple stopping heuristic is met.
- Phase 4: Evaluate quality.

The four-fold stopping heuristic in Phase 2 specifies that screening is terminated only once four independent criteria are met: (1) identification of all predefined relevant records, (2) the number of records screened is at least twice the number of expected relevant records as estimated based on the 1% random records screened as prior knowledge, (3) a minimum of 10% of the dataset is screened, and (4) no relevant records were identified in the last N (e.g. 50) records. In Phase 3 a simple stopping heuristic (e.g., no relevant records in the last N (e.g. 50) records screened) is used.

### 2.3 Study design

To investigate the optimal use of AL-assisted screening tools for screening datasets resulting from broad search strategies, we conducted simulations using four labeled datasets (see Tab. 1) from medication management reviews with ASReview version 1.6.3 [11] (Simulations A-E, G, I, K) and ASReview version 2.0 [33] (Simulations F, H, J, L). Simulation settings are summarized in Tab. 2 and described in more detail below. All scripts and data required to replicate the results are available on Dataverse-NL [34].

**Table 1.** Overview of the datasets. Datasets were obtained by exporting record information and decisions of manual title and abstract screening from the screening tool (Rayyan for datasets 1, 2 and 4; Covidence for dataset 3). FT screening decisions were added manually. TIAB includes = records labeled as (potential) includes during manual title and abstract screening; FT includes = records considered to meet all eligibility criteria and therefore included in the final review. *Excluding 4 FT includes identified in a similar review for a concurrent study [39] and added to the 46 identified within Rayyan.

| Dataset | 1 | 2 | 3 | 4 |
| --- | --- | --- | --- | --- |
| Topic | Prescribing cascades | Adverse drug events in elderly | Pharmacist involvement vs length of stay | Pharmacist technician involvement |
| Search and screening details | [35] | [36] | [37] | [38] |
| # search results | 3475 | 16218 | 6382 | 3878 |
| # TIAB includes | 373 (10%) | 94 (0.6%) | 327 (5%) | 110 (3%) |
| # FT includes | 46* (1%) | 14 (0.08%) | 24 (0.4%) | 20 (0.5%) |
| Screening strategy | dual | single | dual | dual |
| # screeners | 5 | 3 | 3 | 3 |
| reviews | included | excluded | excluded | excluded |

**Table 2.**
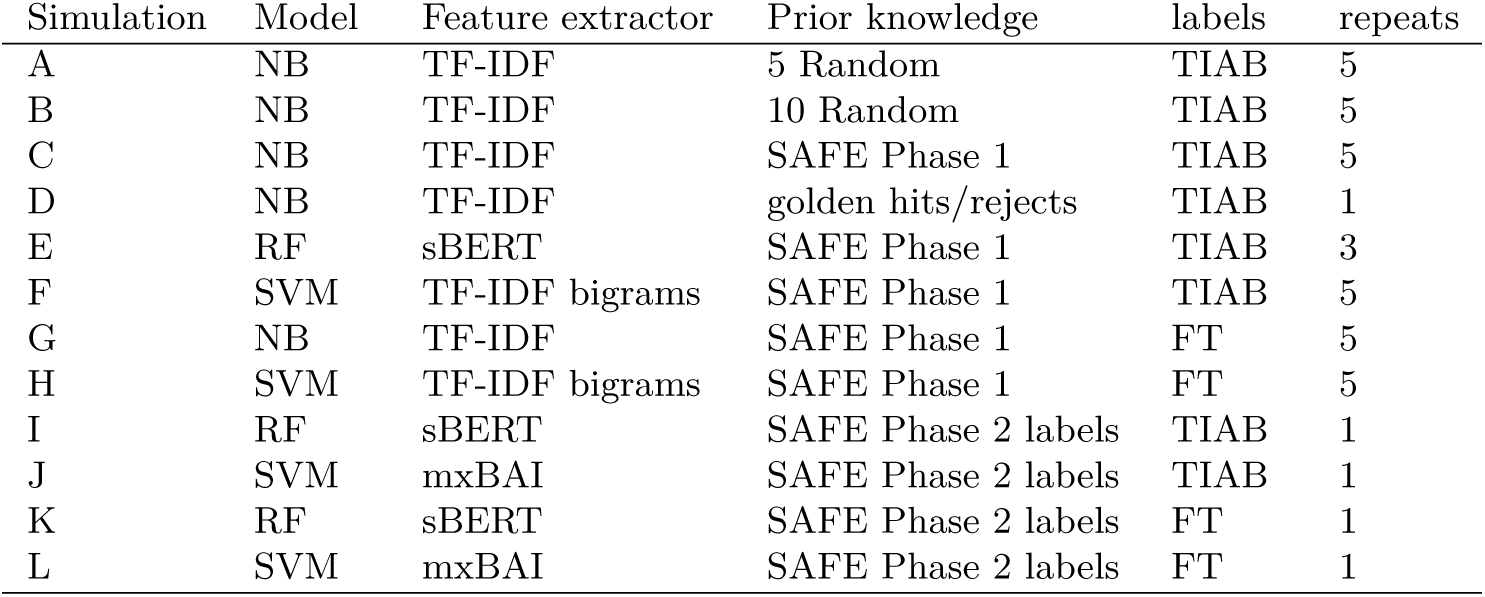
Overview of the choices of intrinsic and extrinsic parameters for all simulations. 5 random = 5 records labeled as include during manual title abstract screening and 5 records labeled as exclude during manual title abstract screening randomly selected from the entire dataset; 10 Random = 10 records labeled as include during manual title abstract screening and 10 records labeled as exclude during manual title abstract screening randomly selected from the entire dataset; golden hits/rejects = 5 records selected by the dataset authors as highly representative of a full text include and 5 records selected by the review authors as highly representative for an absolute exclude during manual title and abstract screening (note that only one such set is available per dataset, hence only one repetition was simulated); SAFE Phase 1 = 1% of data including at least one include; SAFE Phase 2 labels = all records which have labels from Phase 2 (simulations C, F, G or H until SAFE stop heuristic reached for simulations I, J, K or L respectively); TIAB = using include/exclude decisions from manual Title and Abstract screening; FT = using include/exclude decisions from manual full text screening. All simulations use maximum as querying strategy and balanced as balancing approach. Hence the tool-intrinsic parameters of simulations F and H corresponds to ASReview version 2.0’s currently recommended elas-u4 model and those of simulations J and L to ASReview version 2.0’s elas-h3 model.

#### 2.3.1 Datasets

Dataset 1 comes from the published review of Adrien et al. [35]. This review aimed to provide an overview of prescribing cascades that had been identified or confirmed in observational studies. Details on the search strategy and screening process are found in the original publication [35]. Note that during the title/abstract (TIAB) screening stage in Rayyan, topically relevant case reports, case series and reviews were labeled as includes as these were later included in separate forward and backward citation searching to identify relevant publications not retrieved in the systematic search. These records were excluded prior to full text elligibility in post-processing outside Rayyan. In our simulations we use the Rayyan data (373 TIAB includes) instead of the post-processed 249 TIAB includes reported in the PRISMA flowchart in the review. The search results obtained with forward and backward citation searching were not included in our simulations.

The other three datasets are obtained from reviews in progress which were past the full text screening stage.

Dataset 2 originates from a review (in progress) aimed at investigating the short- and long-term health outcomes associated with experiencing an adverse drug reaction or an adverse drug event in older populations. The protocol is available on PROSPERO (CRD42024544258) [36]. At the TIAB screening stage the records were divided among three screeners and each article was evaluated against the eligibility criteria using Rayyan. Articles labeled as potential includes were then assessed by another screener before making a final decision to move an article on to full text screening. At the full text (FT) screening stage eligibility of each article was assessed by two reviewers independently and conflicts were resolved by discussion. During the TIAB screening stage, topically relevant reviews were labeled as excludes.

Dataset 3 originates from a review (in progress) of Holis et al. and the associated published protocol [37]. The review aimed to investigate the effect of hospital pharmacy interventions for patients receiving medical or surgical care on hospital length of stay. Covidence was used for TIAB screening. Each article was independently assessed by two out of three screeners. During the TIAB screening stage, topically relevant reviews were labeled as excludes. This review included peer-reviewed trials with a comparator group.

Dataset 4 originates from a review (in progress) aiming to evaluate the impact of the deployment of pharmaceutical technical assistants on nursing wards. The protocol is available on PROSPERO (CRD42023465217) [38]. Quantitative, experimental studies were included. Roles, responsibilities, and tasks taken up by pharmacy assistants on nursing wards were described. All outcomes of the experimental studies, considered to measure the impact of the deployment of the pharmaceutical technical assistants, for example medication errors, workload, and stock management, were included and summarized in the review. Rayyan was used for TIAB screening. Each article was independently assessed by two out of three screeners.During the TIAB screening stage, topically relevant reviews were labeled as excludes.

#### 2.3.2 Simulation and analysis details

The simulations outlined in Tab. 2 were run for each dataset to assess how various aspects of the SAFE procedure [25] impact on recall and efficiency. The only parameter the SAFE procedure doesn’t provide clear advice for is the labeling strategy. A review of the method sections and supplementary information (SI) of articles citing the use of ASReview in screening (retrieved in Scopus on 29 October 2024) reveals that the tool is used exclusively during title and abstract screening. Hence, when comparing the impact of prior knowledge strategy, choice of model and feature extractor, and stopping heuristics, we used labels from manual title and abstract screening.

Prior to starting the simulations, we assessed whether records erroneously labeled as exclude during manual screening could be identified using ASReview. To do so, ASReview was initialized with a custom Python script that randomly selected 1% of records from each dataset, and also ensured that at least one relevant article was included, in accordance with Step 1 of the SAFE procedure [25] and using the same settings as simulation C. Details of this initialization process are provided in Supplementary Methods 1.1. The ranking of the remaining records was exported. For each dataset, its project lead (FK, CC, TG, TD) evaluated whether records in the top 100 which had previously been excluded at the manual title and abstract screening stage should have been included at the full text stage. For FT includes identified in this step (Tab. 3), the labels (both TIAB and FT) were changed from exclude to include for all simulations A-L.

**Table 3.** AL-assisted screening can help identify full text includes missed by manual screening. For each dataset, the total number of full text includes originally identified using manual screening and the number of extra full text includes identified in the top 100 records considered most likely to be relevant according to ASReview post-initialisation are shown. For the new includes, the ranking according to ASReview is included.

| Dataset | N manual FT includes | # extra FT includes (Ranking ASReview) |
| --- | --- | --- |
| 1 | 46 | 0 |
| 2 | 14 | 1 (31) |
| 3 | 24 | 0 |
| 4 | 20 | 1 (54) |

The SAFE procedure (Phase 1) specifies using minimally 1% of the total number of records (and containing at least one relevant record) as prior knowledge [25]. To compare whether different prior knowledge strategies would lead to significantly different outcomes (measured as recall versus sampling behaviour), four alternative prior knowledge strategies (Simulations A-D) were devised which were performed for each dataset.

For Phase 1 and 2 (training the model with the prior knowledge and the first screening phase), the SAFE procedure advises to using a simple model and feature extractor such as NB and TF-IDF which was also the default setting in ASReview version 1 [25]. This combination was used for Simulations A-D. To evaluate whether a more “complex” initial training model would affect the relevant record retrieval process, Simulation E employed sBERT combined with RF. This approach can demand significantly greater computational resources—particularly with larger datasets—compared to simpler models. Due to the increased processing time and computational load, 3 instead of 5 repetitions were conducted for Simulation E. In the recently released version 2 of ASReview, model elas-u4 is the recommended default [33]. This model uses SVM as classifier and an adapted version of TF-IDF as feature extractor. This choice is tested in Simulation F.

Although it has been suggested that the labeling strategy can significantly impact the performance in AL-assisted screening [24], the SAFE procedure does not provide clear advice on this extrinsic parameter. To assess the extent to which the labeling strategy affects the recall of FT includes over sampling, Simulations C and F were repeated using the FT labels instead of the TIAB labels. These simulations are denoted as Simulations G and H respectively.

The SAFE procedure advises to stop Phase 2 screening once all four of the following criteria are satisfied: 1) at least 10% of the dataset has been screened, 2) all predefined relevant records have been retrieved, 3) the number of records screened is at least twice the number of expected relevant record, and 4) no relevant records were identified in the last N records screened. However, our check of use cases revealed that the simple stopping heuristic “no relevant records identified in the last N records”, with N ranging from 25 to 500 is also commonly used. Therefore, we evaluated the impact of the stopping heuristic on the identification of FT includes and the amount of data that would have been screened for one representative Simulation C and F run. Note that for simulations G and H, where FT labels have been used instead of TIAB labels, only the effect of the SAFE stopping heuristic on recall of FT includes is evaluated.

With our study setup, we could not evaluate the impact of the first criterion of the SAFE stopping heuristic. For all datasets, reaching the third criterion implied having reached the second criterion. Hence for our evaluation, the SAFE stopping heuristic is reduced to a two-fold heuristic requiring that a minimum of 10% of the dataset has been screened and no relevant records were identified in the last N records, where N = 50. This SAFE stopping heuristic is compared to the simple stopping heuristics of encountering N = 50, 100, or 200 consecutive irrelevant records.

Once the stopping heuristic is reached, the SAFE procedure requires a third phase where the decisions (both prior knowledge and Phase 2 screening) prior to reaching the stopping heuristic should be used as prior knowledge to train a more advanced model and feature extractor combination, which are then used to continue screening until again no relevant records are encountered in the last N records screened (Phase 3) [25].

Whether or not it is necessary and useful to carry out Phase 3 of the SAFE procedure depends on two things: 1) whether or not all final full text includes would have been retrieved in Phase 2 before the stopping heuristic was met and 2) on whether or not the remaining full text includes are identified in Phase 3 before the stopping heuristic of Phase 3 is reached. To assess this using our manually labeled datasets, for one representative Simulation C, F, G and H run per dataset, we determined the number of full text includes identified before meeting the stopping heuristic. To assess the extra workload associated with this step, we also evaluated the extra amount of data which would have been screened before reaching the stopping heuristic for Phase 3.

Subsequently for all datasets four “complex simulations” were run. Simulations I and K used all labeling decisions from respectively the analyzed Simulation C or G run up till the SAFE stopping heuristic was reached as prior knowledge followed by simulated screening with Random Forests and sBERT as classifier and feature extractor. Simulations J and L used all labeling decisions from respectively the analyzed Simulation F or H run up till the SAFE stopping heuristic was reached as prior knowledge followed by simulated screening with SVM and mxBAI-embed-large-v1 as model and classifier (elas-h3 in ASReview 2.0). The resulting Phase 3 simulations were assessed for their ability to capture remaining FT includes before reaching the stopping heuristic of N=50 consecutive irrelevant records and how much extra screening this would cost compared to stopping after Phase 2.

## 3 Results

### 3.1 Can AL-assisted screening aid in identifying records missed in manual screening?

As summarized in Table 3, for two of our four datasets an extra full text include was discovered when reviewing the top 100 articles after initialisation of ASReview following the SAFE procedure.

### 3.2 Is the SAFE procedure sufficient, necessary, and efficient when using AL-assisted screening?

We assessed the recommendations laid out in the SAFE procedure for their impact on recall and efficiency.

When applying the various stopping heuristics to the original SAFE Phase 2 simulation (Simulation C), only N=200 consecutive irrelevant results achieves 100% recall of full text includes across all four datasets. However, this choice comes at the price of severely decreased efficiency, particularly for datasets 1 and 3. When applying the stopping heuristics to the newly recommended elas-u4 model (Simulation F), none achieve 100% recall of full text includes. For both the original and the new model, the SAFE stopping heuristic results in recall of at least 93% across datasets. For our datasets this means that at most one full text include is missed when applying this stopping heuristic. Applying the criteria of N=50 or N=100 irrelevant records in a row shows inconsistent recall and efficiency behavior across datasets. Overall, the SAFE Phase 2 stopping heuristic strikes a good balance between recall and efficiency and performs similarly across datasets. When applying this stopping heuristic the elas-u4 option showed improved efficiency for datasets 3 and 4 but decreased efficiency for datasets 1 and 2.

Despite there being evidence that the labeling strategy employed by the user can have a significant impact on the performance of AL-assisted screening [24], the SAFE procedure provides no clear advice on this tool-extrinsic parameter. We evaluated the impact of using labels from manual full text screening decisions (Simulations G and H) versus those from manual title-abstract screening (Simulations C and F respectively) on the recall versus sampling behavior (Fig. 2 and SI Fig. 3). Comparing the recall versus sampling plots (Fig. 2) and the percentage of data screened at discovery of the final FT include (Tab. 4) of simulations C and G or F and H makes it clear that using FT labels instead of TIAB labels significantly improves recall over sampling for datasets 1, 3 and 4. When using the NB and TF-IDF combination (Simulations C, G) the final full text include in these datasets would have been discovered after screening 15-18% of the dataset if FT labeling is used compared to after screening 18-32% of the dataset using TIAB labeling. When using the new elas-u4 combination, dataset 2 shows notably different behaviour from the other datasets. For datasets 1, 3 and 4 using FT labels instead of TIAB labels results in highly improved recall over sampling with the final FT includes being identified within screening 7-11% of the dataset (compared to 13-27% when using TIAB labels). For dataset 2, which has a very low number of full text includes relative to the entire dataset, switching to FT labels decreases recall versus sampling. when FT labels are used 23% of the dataset needs to be screened to identify the final FT include whereas the final FT include would have been identified after screening 15% of the dataset if TIAB labels would have been used.

**Table 4:** Impact of stopping heuristic and labeling strategy on retrieval of FT includes and amount of data to be screened. Stop. Heur. = the stopping heuristic applied; SAFE2 = 10% of the dataset and N=50 irrelevant records in a row; N=50, 100, 200 refers to encountering 50, 100, 200 irrelevant records in a row; SAFE3 = the recommended stopping heuristic for SAFE protocol Phase 3 (N=50); Last FT = how much of the dataset would have to be screened to retrieve the last FT include in this simulation; Screen stop = part of the dataset that has been screened when the stopping heuristic is satisfied; FT retrieved = part of the full text includes retrieved when the stopping heuristic is satisfied. n.a. = all FT includes were already discovered in earlier phases.

| Dataset | Simulation | Stop. Heur. | Last FT | Screen stop | FT retrieved |
| --- | --- | --- | --- | --- | --- |
| 1 | C | SAFE2 | 27% | 20% | 98% |
| 1 | C | N=50 | 27% | 20% | 98% |
| 1 | C | N=100 | 27% | 87% | 100% |
| 1 | C | N=200 | 27% | 100% | 100% |
| 1 | F | SAFE2 | 27% | 24% | 98% |
| 1 | F | N=50 | 27% | 24% | 98% |
| 1 | F | N=100 | 27% | 73% | 100% |
| 1 | F | N=200 | 27% | 92% | 100% |
| 1 | G | SAFE2 | 16% | 10% | 93% |
| 1 | H | SAFE2 | 11% | 10% | 98% |
| 1 | I | SAFE3 | 22% | 55% | 100% |
| 1 | J | SAFE3 | 26% | 30% | 100% |
| 1 | K | SAFE3 | 15% | 14% | 96% |
| 1 | L | SAFE3 | 11% | 11% | 100% |
| 2 | C | SAFE2 | 8% | 10% | 100% |
| 2 | C | N=50 | 8% | 3% | 67% |
| 2 | C | N=100 | 8% | 7% | 93% |
| 2 | C | N=200 | 8% | 9% | 100% |
| 2 | F | SAFE2 | 15% | 10% | 93% |
| 2 | F | N=50 | 15% | 4% | 67% |
| 2 | F | N=100 | 15% | 7% | 87% |
| 2 | F | N=200 | 15% | 8% | 87% |
| 2 | G | SAFE2 | 73% | 10% | 73% |
| 2 | H | SAFE2 | 23% | 10% | 87% |
| 2 | I | SAFE3 | n.a. | 10% | 100% |
| 2 | J | SAFE3 | 12% | 11% | 93% |
| 2 | K | SAFE3 | 33% | 10% | 80% |
| 2 | L | SAFE3 | 16% | 10% | 87% |
| 3 | C | SAFE2 | 32% | 16% | 83% |
| 3 | C | N=50 | 32% | 16% | 83% |
| 3 | C | N=100 | 32% | 32% | 96% |
| 3 | C | N=200 | 32% | 50% | 100% |
| 3 | F | SAFE2 | 26% | 13% | 88% |
| 3 | F | N=50 | 26% | 13% | 88% |
| 3 | F | N=100 | 26% | 28% | 100% |
| 3 | F | N=200 | 26% | 30% | 100% |
| 3 | G | SAFE2 | 20% | 10% | 96% |
| 3 | H | SAFE2 | 7% | 10% | 100% |
| 3 | I | SAFE3 | 34% | 30% | 96% |
| 3 | J | SAFE3 | 27% | 20% | 92% |
| 3 | K | SAFE3 | 20% | 11% | 96% |
| 3 | L | SAFE3 | n.a. | 10% | 100% |
| 4 | C | SAFE2 | 18% | 14% | 95% |
| 4 | C | N=50 | 18% | 14% | 95% |
| 4 | C | N=100 | 18% | 29% | 100% |
| 4 | C | N=200 | 18% | 32% | 100% |
| 4 | F | SAFE2 | 13% | 18% | 100% |
| 4 | F | N=50 | 13% | 18% | 100% |
| 4 | F | N=100 | 13% | 23% | 100% |
| 4 | F | N=200 | 13% | 33% | 100% |
| 4 | G | SAFE2 | 15% | 10% | 86% |
| 4 | H | SAFE2 | 9% | 10% | 100% |
| 4 | I | SAFE3 | 20% | 20% | 95% |
| 4 | J | SAFE3 | n.a. | 24% | 100% |
| 4 | K | SAFE3 | 47% | 12% | 86% |
| 4 | L | SAFE3 | n.a. | 10% | 100% |

Applying the standard SAFE Phase 2 stopping heuristic in combination with FT labeling (Tab. 4, Simulations G, H) results in termination after screening 10% of the dataset for all four datasets. If screening is simulated with the NB and TF-IDF combination (Simulation G), this causes a decreased identification of FT includes for datasets 2, 3, and 4, despite this setup’s improved efficiency at identifying FT includes. This indicates that this stopping heuristic may not be appropriate to use in combination with FT labeling. When screening is simulated with elas-u4 (Simulation H), only dataset 2 shows decreased FT include retrieval which is in line with the decreased recall versus sampling performance observed.

We evaluated whether the recommended second screening phase (Phase 3) of the SAFE procedure is capable of identifying FT includes which were not identified in Phase 2 prior to reaching the stopping heuristic (Simulations I-L, SI Fig. 4 and SI Fig. 5). For all combinations of labeling strategy, model and feature extractor tested, Phase 3 captured some of remaining FT includes for some (not all) of the datasets. Two combinations achieve 100% recall of FT includes: the original Naive Bayes with TF-IDF for Phase 1 and 2 followed by sBERT with RF for Phase 3 and using TIAB labels and the elas-u4 for Phase 1 and 2 followed by elas-h3 for phase 3 and FT labels. The first combination only misses 1 FT include for dataset 3. The second combination misses 2 FT includes for dataset 2. Out of the two approaches, the latter is far more efficient. Only 10-11% of the dataset would have been screened whereas 10-55% of the dataset would have been screened with the first approach.

## 4 Discussion

Our results confirm that AL-assisted screening has the potential to greatly reduce the workload in screening the large, heterogeneous datasets common in medication management reviews whilst maintaining a recall rate of FT includes similar to full manual screening.

Our results indicate that overall the SAFE procedure [25] strikes a good balance between efficiency and recall (Tab. 4). For three of our datasets, the best performance is achieved with the combination of elas-u4 (Phase 1 and 2), elas-h3 (Phase 3) available in ASReview version 2 [33] and using FT labels (in combination with the original SAFE prior knowledge strategy and stopping heuristics (Phase 2 and 3)). This combination resulted in 100% recall of FT includes and screening only 10-11% of the dataset. This choice of parameters, however, does not perform well for dataset 2. For dataset 2, the best performance is achieved with the combination of NB, TF-IDF and TIAB labeling. Using this combination, all FT includes are retrieved in Phase 2 upon reaching the SAFE Phase 2 stopping criteria after screening 10% of the dataset.

Dataset 2 differs from the other datasets in that it is significantly larger and has a very small percentage of TIAB and, especially, FT includes (see Tab. 1). It is likely that the simplest model and feature extractor combination are better suited at dealing with such extremely unbalanced datasets. This would also explain why the combination of sBERT with RF (Simulation E) shows worse recall versus sampling for dataset 2 only. Moreover, the number of FT includes may simply be too low for even the simple models to learn about the characteristics of an include.

Previous validation studies have highlighted that the active learning model choice, and in particularly the feature extractor can affect the discovery times of relevant records [20, 22]. Several studies suggest that the relatively simple TF-IDF feature extractor outperforms other techniques in work saved over sampling, time to discovery, and recall [21, 22]. It seems that simpler models, such as Näıve Bayes or Logistic Regression, outperform more advanced models when limited training data is available, such as in the early phases of screening and for smaller datasets.

Based on this, we would have expected a larger negative impact of choosing sBERT with RF on the recall versus sampling behavior (Fig. 1). However, it is not unlikely that the expected loss in performance is (partially) offset by the improved ability of this advanced combination to deal with relatively heterogeneous datasets obtained by broad search strategies likes the ones used for our reviews. Our results indicate elas-u4 model and feature extractor combination advised as the default choice in ASReview version 2 [33] combines the best of both worlds, showing improved recall versus sampling behavior for all datasets and labeling strategies.

**Fig. 1.**
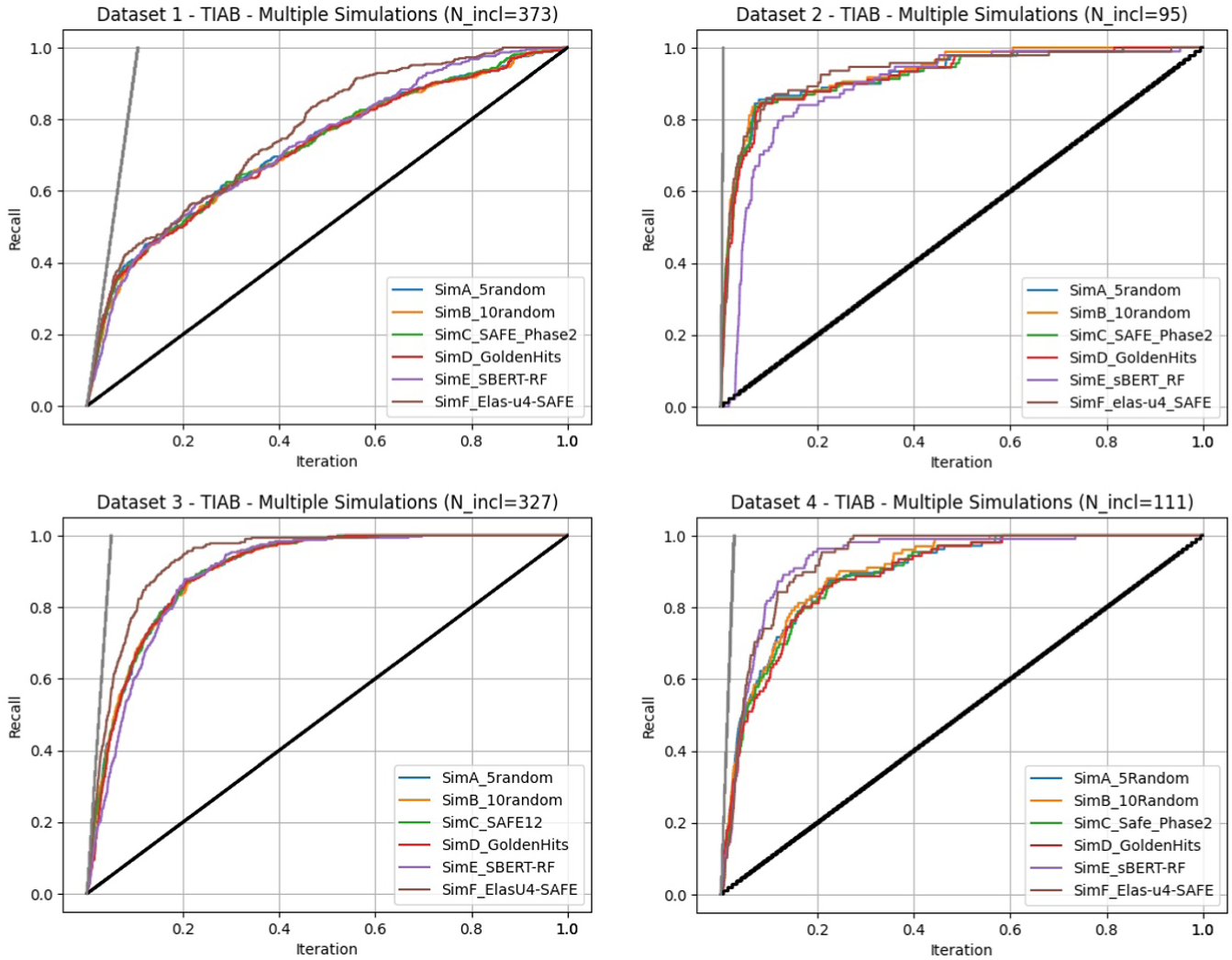
Impact of prior knowledge strategy and model and feature extractor choice on recall over sampling. Simulations A-F for each dataset separately. Simulations A-D for each dataset show how recall versus sampling depends on prior knowledge strategy. Simulations C, E and F show how the choices for model and feature extractor affect recall versus sampling for each dataset. Details of the simulations can be found in Tab. 2. The black lines indicate recall versus sampling when the dataset would have been sampled at random. All runs for the simulations related to prior knowledge strategy are shown in SI Fig. 1 and those related to model and feature extractor selection in SI Fig. 2.

When using TIAB labels, we observed that the recall versus sampling behavior is very dataset dependent (Fig. 1). This may be due to heterogeneity in the search results, heterogeneity in the articles which cannot confidently be excluded in the TIAB screening stage, or both. When using FT labels, the recall versus sampling behavior we observed is much less dataset dependent (Fig. 2).

**Fig. 2.**
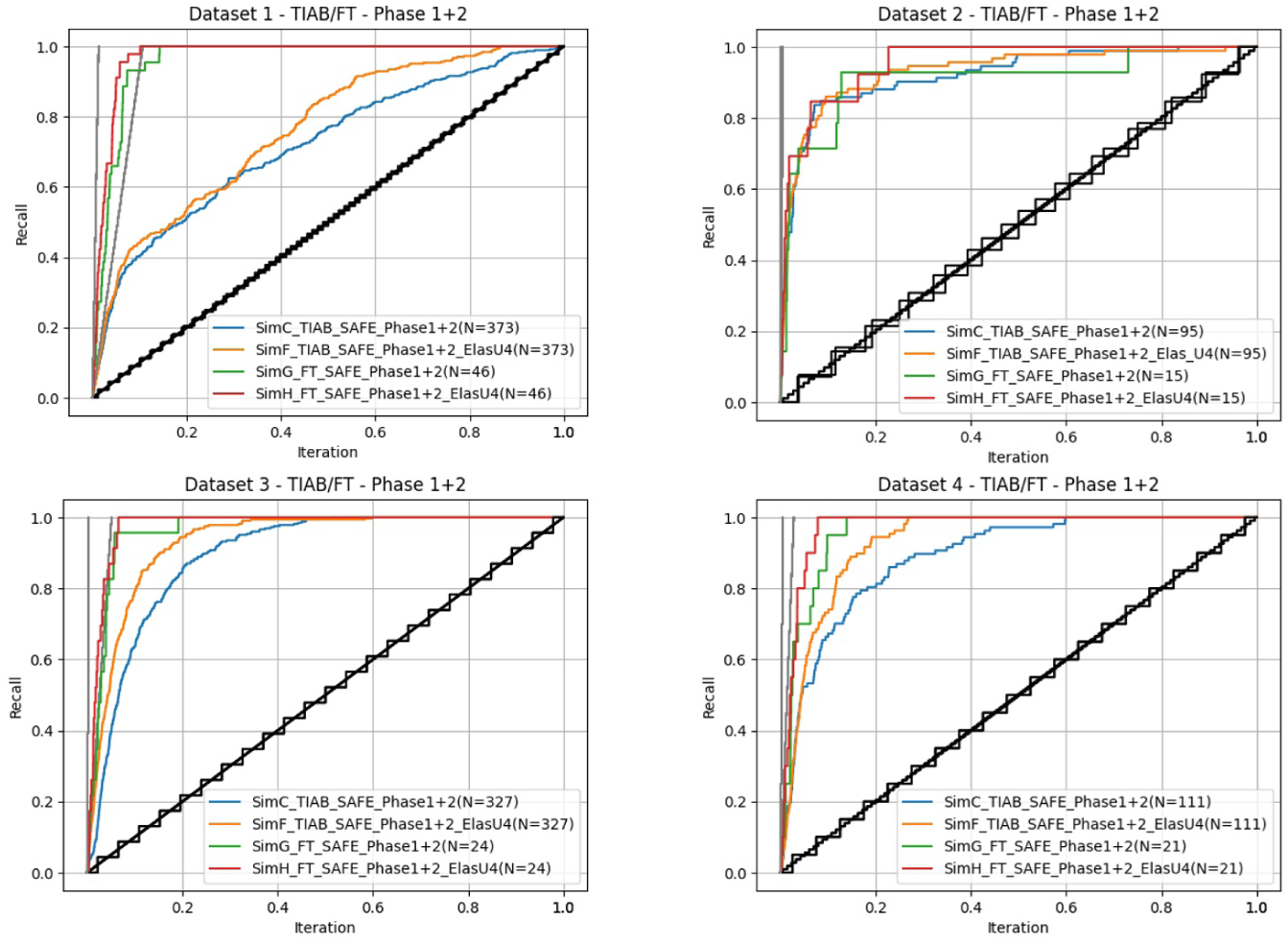
Impact of labeling strategy on recall versus sampling. Recall versus sampling plots for simulations G (blue) and H (green) using the FT labeling decisions instead of the TIAB labeling decisions for each dataset separately. The black lines indicate recall versus sampling when the dataset would have been sampled at random. For comparison, the recall versus sampling plots for the corresponding TIAB labeling simulations are shown as well (Simulation C in red, F in orange).

It seems intuitive that noisier labels may negatively impact the performance of AL-assisted screening. In other words, if records are often incorrectly labeled as “include”, the model will have a fuzzier (noisier) understanding of the characteristics of an include and is more likely to prioritize the wrong records in the ranking. Noisy labeling may result from inconsistent application of the predefined eligibility criteria, articles simply not providing sufficient detail to be excluded, or the choice to include source types (e.g. reviews to be used for subsequent snowballing) at the TIAB screening stage which do not match the inclusion criteria. Labeling decision made in title-abstract screening are also by definition noisier than those made in full text screening.

A recent study focused on searches for medical guideline development indeed confirms that performance of AL-assisted screening depends on how noisy the labeling decisions are [24]. The datasets used in this study are derived from highly specific searches and are comparatively small (56-772 records). Our results confirm this observation also holds for large, more heterogeneous datasets resulting from broad search strategies in the field of medication management (Fig. 2). It is interesting to note here that we used the TIAB labels post conflict resolution. Prior to conflict resolution, the TIAB labels are likely to be even noisier, potentially making the effect of switching to FT labels even larger when using AL-assisted screening.

We only compared the SAFE Phase 2 stopping heuristic to simpler but popular stopping heuristics in combination with using a TIAB labeling strategy (Simulations C and F). A simple stopping heuristic of encountering N irrelevant results in a row would not work well in combination with FT labeling due to the very low percentage of FT includes in the datasets.

The SAFE stopping heuristic showed more consistent behavior across datasets than using a simple stopping heuristic based only on the number of consecutive irrelevant records. N = 50 or 100 led to decreased recall of FT includes for some datasets and N = 100 or 200 negatively impacted efficiency in some datasets. By contrast, the SAFE stopping heuristic struck a good balance between efficiency and recall for all datasets.

It should be noted that the SAFE stopping heuristic in most cases does not result in 100% recall of FT includes. Continuing with Phase 3, as recommended, results in improved recall. When using the elas-u4 (Phase 1 and 2) followed by elas-h3 (Phase 3), the extra screening burden due to this is low.

Although selecting prior knowledge is essential to start AL-assisted screening, it has been suggested that performance of AL-assisted screening is relatively insensitive to the choice of prior knowledge [20]. Similarly, another study reports that no clear trends could be established across datasets [21]. Our results confirm that the recall versus sampling behavior is relatively insensitive to the prior knowledge strategy (Fig. 1).

Although this SAFE prior knowledge strategy does not significantly improve recall versus sampling, it offers one important benefit: one can estimate the total number of relevant records in the entire dataset.

Note that, depending on the labeling strategy, the chances of not encountering any relevant records in the randomly selected 1% can be high. For our datasets, the ratio of relevant records to total records ranges from 0.6-10% when using TIAB labels. This reduces to 0.09-1% when FT labels are used instead. Given that any AL-assisted screening tool will need at least one example of a relevant record and that the performance seems rather insensitive to the prior knowledge strategy applied, we recommend to manually add a pre-known relevant record present in the dataset to the prior knowledge set of the 1% selected at random if no relevant records were included. The estimate of relevant records in the dataset can then be calculated using standard statistics.

Even review teams preferring to rely on manual screening can benefit from AL-assisted screening tools. We have shown that AL-assisted screening can help identify relevant records that were wrongly dismissed during manual TIAB screening (Table 3). Hence, similar to our test here, a team could complete full screening as normal and subsequently initialize ASReview according to the SAFE procedure using random sampling from their labeled dataset, and screen through the resulting top 100 or 200 records. Alternatively, an additional screener could screen using AL-assisted screening and compare their results to those of the manual screeners during conflict resolution. However, our results support replacing manual screening with AL-assisted screening as a means to reduce time spent on screening irrelevant records whilst maintaining an error rate comparable to that of the gold standard of manual dual screening.

Note that the number of (tool-intrinsic and tool-extrinsic) parameters which should be considered when using AL-assisted screening may seem intimidating. However, new versions of existing AL-assisted screening tools such as ASReview version 2 [33] have improved user interfaces and are similarly easy to set up and use as commonly used sreening tools such as Covidence or Rayyan.

### 4.1 Limitations of this study and further research

Our study focuses on the application of AL-assisted screening in medication management reviews. Such reviews typically rely on broad, unspecific search strategies which yield large, heterogeneous datasets. The generalizability of our results to other fields where such datasets are common should be verified in future work.

In our simulations, we used the TIAB labels post conflict resolution. As there can be significant mismatch between reviewers [4, 5, 40], it would be interesting to see how the differences between reviewers affect discovery of relevant records.

In this study we focused on AL-assisted screening but this is not the only method by which large language models can potentially aid researchers in lowering the number of articles to screen [12, 41, 42]. Recently a comparison of AL-assisted screening and prompt-based screening with a large language model for datasets of review in thoracic surgery indicated that the latter may even outperform AL-assisted screening in efficiency gain [43]. Future research should explore whether this observations holds for reviews in other fields, including those with broader search strategies, such that an optimal workflow can be developed for researchers.

## 5 Conclusions

Using AL-assisted screening instead of, or in addition to, manual screening can significantly reduce the number of records needed to screen whilst achieving a recall of FT includes comparable to or better than that obtained with full manual screening in medication management systematic reviews. The optimal choice of parameters when using AL-assisted screening depends on the dataset characteristics. To screen the large and heterogeneous datasets resulting from broad search strategies, in most cases we recommend a modified version of the SAFE procedure where FT labeling is used instead of TIAB labeling in combination with the elas-u4 (Phase 1 and 2) and elas-h3 (Phase 3) models available in ASReview version 2. Should a dataset have a very low percentage of TIAB and FT includes (e.g. lower than 2% and 0.2% respectively), using the simpler Naive Bayes and TF-IDF combination and TIAB labeling may be more appropriate. These numbers can be estimated from the prior knowledge strategy where 1% of the data is sampled at random.

## Data Availability

All data produced in the present study are available upon publication of the mentioned datasets, scripts, code, and their results used in this study will be made openly available after the associated datasets have been released/published.

## Supplementary information

The supplementary information (SI) contains more detailed information on the methods and the recall versus sampling plots for all simulation runs.

## Acknowledgements

We thank Roberto Cruz Martinez and Floor Ruiter for their feedback on the study design, interpretation of results, and the manuscript.

## Declarations

- Funding Not applicable.
- Conflict of interest/Competing interests. The authors declare that they have no conflicts of interest.
- Ethics approval and consent to participate. Not applicable.
- Consent for publication. Not applicable.
- Data availability. The datasets used in this study are derived from ongoing or forthcoming systematic reviews. To respect publication policies of those primary review manuscripts, the datasets cannot be shared before the corresponding reviews are published. After publication of the source reviews, all datasets will be made openly available at Dataverse-NL [34]
- Code availability. All scripts, code, and their results used in this study will be made openly available at Dataverse-NL [34] after the associated datasets have been released/published.
- Author contribution. Conceptualization: MS, FKC, RGV. Methodology: MS, RGV. Software & Formal analysis: RGV. Data curation: MDG, RVH, JF, PD, TG, FKC, TD, CC. Writing: Original draft: MS, RGV, MDG, FKC, TD, CC, TG. Writing: Review & Editing: All authors.

