## Supplementary Information for "To Include or Not to Include? A prescription from the pharmacy on how to use active learning assisted screening in systematic reviews"

Verdonschot, R.G, et al. (2025)

### 1 Supplementary Methods

To ensure reproducibility of the results, ASReview simulations allow for the selection of two seed numbers called `seed` and `init_seed`. `Seed` specifies the seed for the random number generator used in active learning during the simulation, ensuring that the order in which records are sampled (i.e., suggested for labeling) remains consistent across runs and can be replicated. `Init_Seed` specifies the seed for the random number generator used to select the initial set of labeled records, ensuring the same initial subset of records is used to "warm-up" the simulation.

For Simulations A and B the seed numbers were selected randomly through a website called https://www.random.org/. The following example shows a Windows powershell command used in Simulations A and B (whereby seeds 173 and 29 were obtained using random.org). Each simulation obviously had different seed numbers. All scripts are available through our Dataverse repository.

```
asreview simulate Case1.xlsx -s Case1_SimA_Run1.asreview
    --n_prior_included 5 --n_prior_excluded 5 --seed 173
    --init_seed 29
```

Simulations C and E-H were performed using a custom python script (see the SI and available at the Dataverse repository) in which the seed was obtained using Python's `random` package. Simulation D was performed without seeding for the selection of prior knowledge as this was Golden Hits and Rejects are handpicked.

#### 1.1 How are priors selected in SAFE Phase 1?

All scripts and data can be found on the following repository (https://doi.org/10.34894/OZYH98/) after the associated datasets have been released. To initialize each simulation using SAFE Phase 1, a python script selects a random subset of 1% of the dataset (rounded up). This is done using:

`num_to_select = math.ceil(len(df) * 0.01)`

Then, a random sample of that particular size (`num_to_select`) is drawn from the dataset with `random.sample`, and the number of relevant articles (i.e., those labeled as `Included = 1`) is counted using:

`included_count = selected_articles['Included'].sum()`

To ensure that each simulation begins with at least one relevant article, the script uses a `while` loop that continues sampling until this condition is satisfied:

```
while True:
    selected_articles, included_count = select_articles()
    if included_count >= 1:
        break
```

This guarantees that every simulation adheres to the SAFE procedure by starting with a minimally informative prior set (of 1%) containing at least one relevant study.

### 2 Supplementary Figures

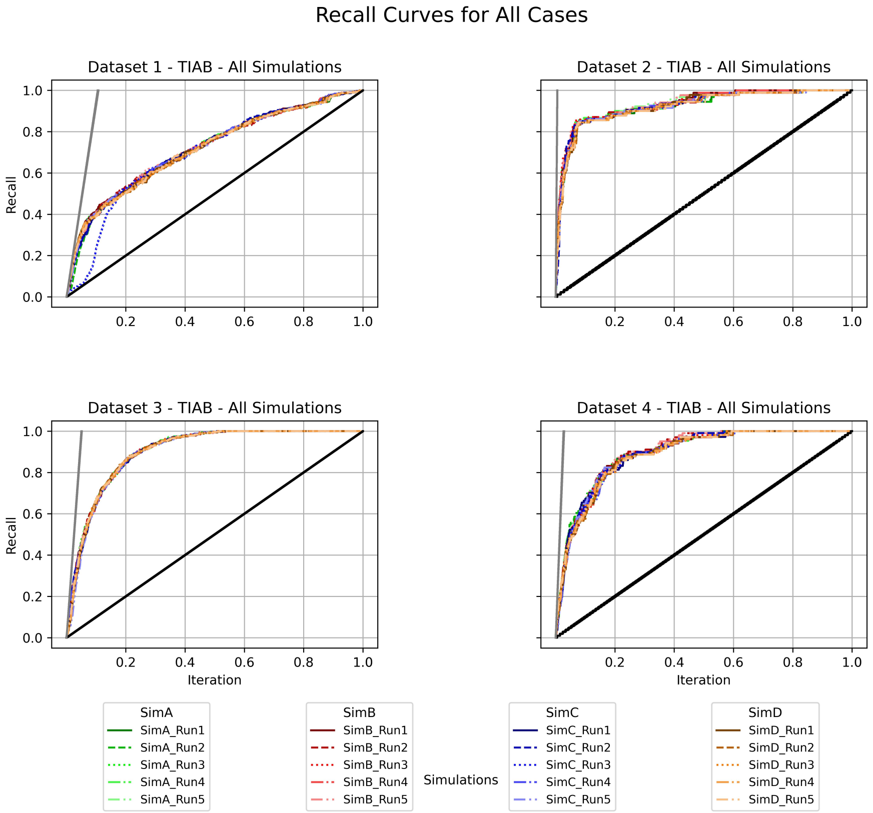


**Fig. 1** Supplementary Figure 1: Comparing prior knowledge strategies. For each dataset, all runs of simulations A-D are shown

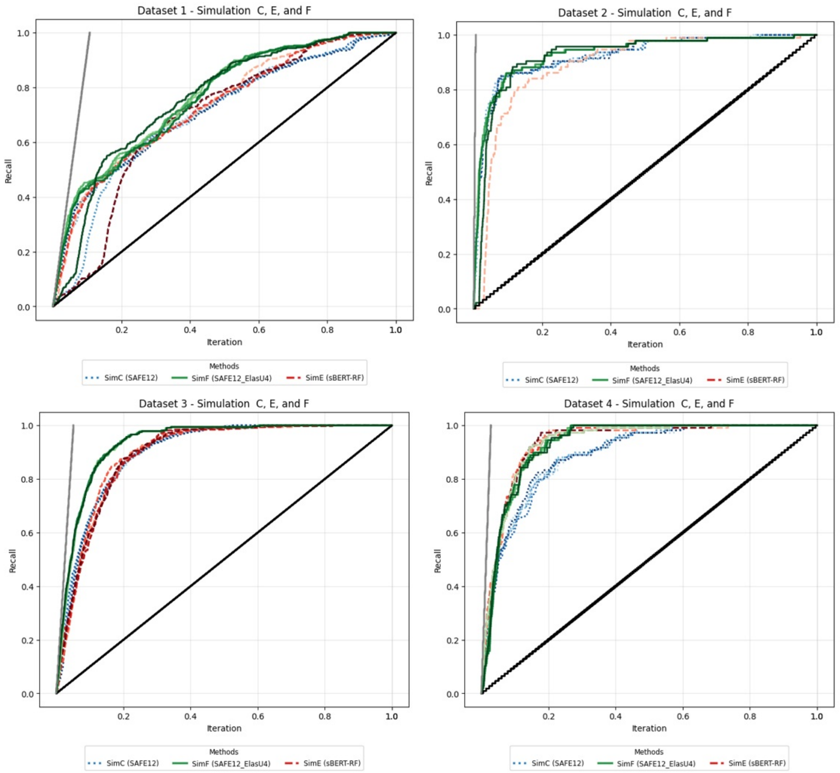


**Fig. 2** Supplementary Figure 2: Comparing choice of model and feature extractor. For each dataset, all runs of simulations C,E and F are shown

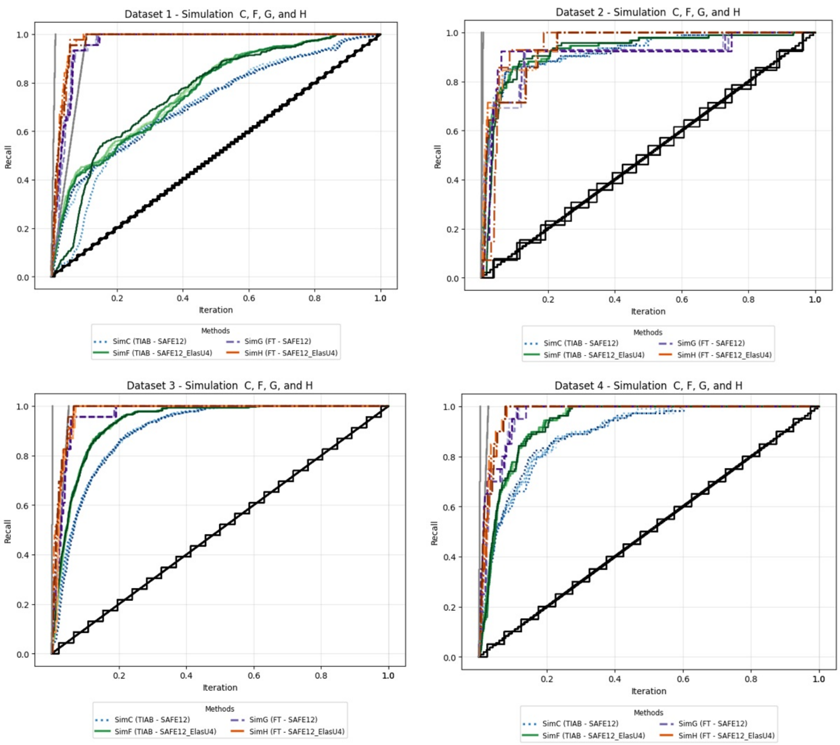


**Fig. 3** Supplementary Figure 3: Comparing labeling strategies. For each dataset, all runs of simulations C, F, G, H are shown

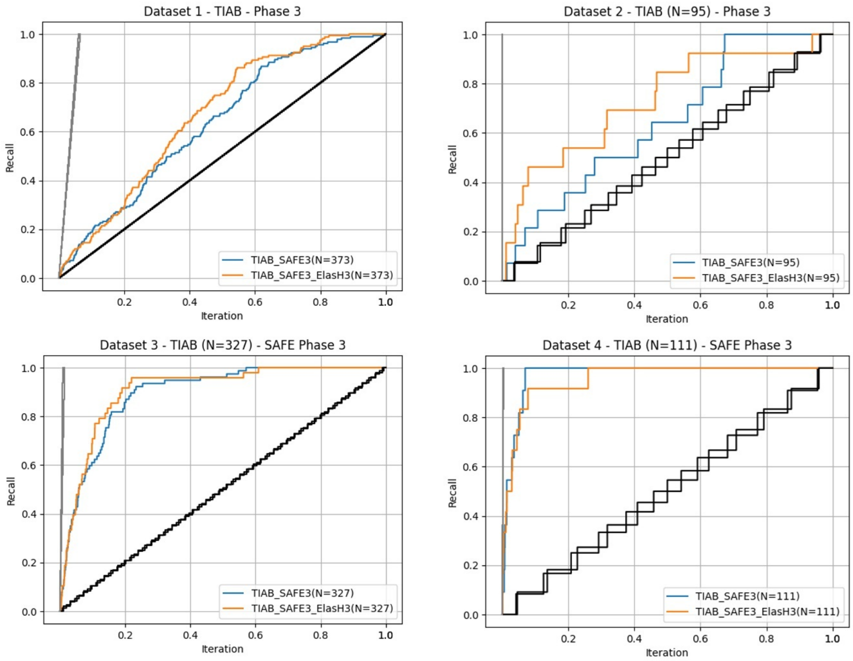


**Fig. 4** Supplementary Figure 4: SAFE Phase 3 recall versus sampling using TIAB labels. For each dataset, recall versus sampling of simulations I and J are shown

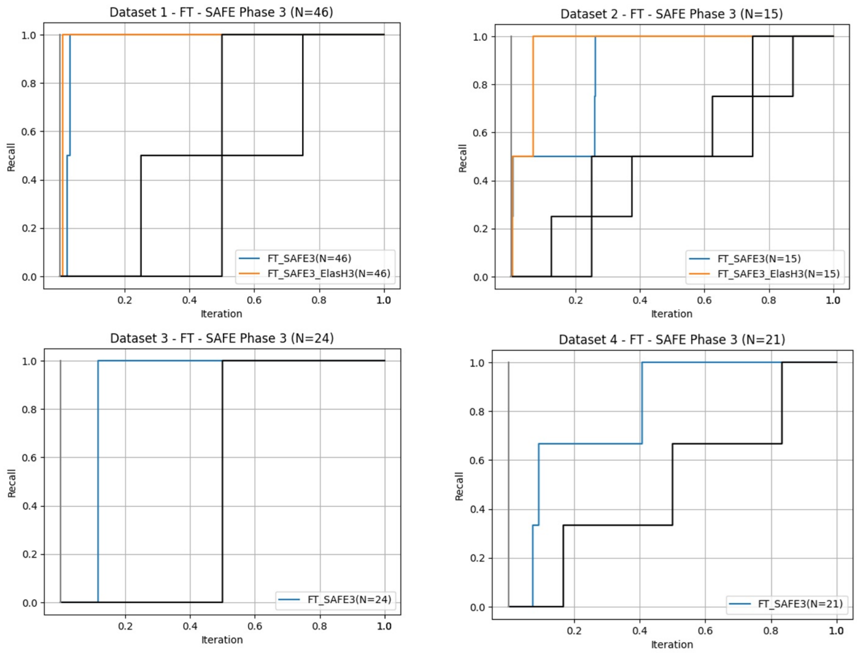


**Fig. 5** Supplementary Figure 5: SAFE Phase 3 recall versus sampling using FT labels. For each dataset, recall versus sampling of simulations K and L are shown. Note that simulation L was not ran for datasets 3 and 4 as no FT includes were left in the dataset post SAFE Phase 2.
